# Repetitive negative thinking is associated with amyloid accumulation over 3 years in cognitively healthy adults

**DOI:** 10.64898/2026.10.29.26366526

**Authors:** Natalie L. Marchant, Florence Mezenge, Anjali Raghavan, Elise Saul, Vincent de la Sayette, The IMAP research group, Gaël Chételat

**Affiliations:** Division of Psychiatry, University College London, London, United Kingdom; Normandie Univ, UNICAEN, INSERM, UA20, NEUROPRESAGE, Cyceron, Caen, France; CHU Caen-Normandie, Department of Clinical Research, Caen, 14000, France

## Abstract

**Importance:** Identifying potentially modifiable risk factors for Alzheimer’s disease (AD) is a priority. Repetitive negative thinking (RNT) is a modifiable cognitive-emotional process common to many psychological risk factors for AD, including anxiety and depression. The psychological debt hypothesis proposes that RNT may itself increase risk for AD.

**Objective:** To determine whether RNT symptoms are associated with change in established neurobiological markers of AD risk over time.

**Design:** Longitudinal neuroimaging study with the IMAP/IMAP+ cohort.

**Setting:** Monocentric with community-dwelling participants.

**Participants:** 94 cognitively healthy adults aged 21-89 years (average 57.41 SD 18.31, 55% female) with up to 3 neuroimaging scans over a 3-year period.

**Exposure:** All participants completed validated measures of RNT, anxiety and depression symptoms.

**Main Outcome(s) and Measure(s):** Change over time in global cortical amyloid (Florbetapir-PET), glucose metabolism (FDG-PET), and structural grey matter volume (MRI) adjusted for potential confounders. PET images were corrected for partial volume effects, and AD-relevant masks were used for all neuroimaging outcomes. Post-hoc analyses divided the predictor variable into tertiles to aid visualisation, and FDR-corrected voxel-wise analyses were performed to identify region-specific changes.

**Results:** RNT was associated with increasing global amyloid accumulation even after controlling for psychological symptoms (*B* = 0.0120, 95% Confidence Interval 0.003 to 0.021, *P* = 0.012), and in sensitivity analyses limited to participants aged ≥50 or ≥60. Post-hoc tests showed a significant difference in rate of amyloid accumulation between the lowest and highest RNT tertiles (*P*=0.008). Voxelwise analyses revealed that RNT predicted significantly faster increases in regions vulnerable to early amyloid accumulation in AD (e.g. precuneus, posterior cingulate cortex, and medial frontal cortex). RNT was not associated with change in glucose metabolism or brain atrophy (*P*s>0.05). Neither anxiety nor depressive symptoms were associated with change in any neuroimaging outcome (*P*s>0.05).

**Conclusions and Relevance:** Higher RNT was associated with more rapid accumulation of cortical amyloid in regions affected early in AD. These findings indicate that this transdiagnostic cognitive-emotional process may be an early marker and/or driver of AD pathology. Given that RNT is responsive to treatment, future research is needed to investigate whether modifying RNT reduces AD risk.

## INTRODUCTION

Alzheimer’s Disease (AD) is a progressive neurodegenerative disease characterised by accumulation of β-Amyloid (Aβ) and hyperphosphorylated tau proteins and atrophy in the brain.^1^ In the absence of established disease-modifying treatments, there is an urgent need to identify modifiable risk factors that are associated with changes in these biomarkers, which can be targeted to prevent future AD dementia. These changes precede cognitive impairment,^2^ allowing us to examine premorbid associations between risk factors and neuropathological change.

In recent decades, a number of psychological risk factors for cognitive decline and AD have been identified, which include depression^3^ and anxiety.^4^ While these risks have generally been considered independently, the psychological debt (formerly termed cognitive debt) hypothesis suggests that a mechanism frequently present in these psychological factors—repetitive negative thinking (RNT) —may underlie the risk associated with each factor.^5,6^ RNT is a modifiable, transdiagnostic process that encompasses both future-directed worry and past-directed ruminative brooding.^7^

Accumulating evidence supports this hypothesis. Higher levels of RNT have been associated with subjective^8,9^ and objective cognition (in samples with lower education),^10^ and more rapid decline in objective cognitive domains affected early in AD dementia, specifically global cognition, immediate and delayed memory.^11,12^ While no study has yet investigated its direct link with incident dementia, worry, examined as a nuance of neuroticism, was specifically associated with elevated dementia risk.^13^ Further, higher levels of RNT have been cross-sectionally associated with amyloid,^11,14,15^ (but not always^16^) neurofilament light chain in women (an indicator of neurodegeneration),^17^ greater brain age,^18^ and altered functional connectivity within brain networks associated with AD.^19^ Further research is needed to establish whether RNT is associated with *change* in neuropathological AD markers in cognitively unimpaired individuals to ascertain whether it could be a marker of early pathological processes and serve as a potential target for prevention.

Here we sought to determine whether RNT is associated with longitudinal change in established neurobiological markers of AD, specifically neuroimaging measures of amyloid, glucose metabolism, and grey matter volume. We secondarily examined whether anxiety or depressive symptoms were associated with changes in these markers.

## METHODS

### Participants

94 cognitively healthy adults were included from the multimodal neuroimaging study of early AD (Imagerie Multimodale de la maladie d’ Alzheimer à un stade Précoce [IMAP/IMAP+]) which began in 2008 in Caen, France. Eligibility criteria are detailed in previous publications.^20–22^ Briefly, participants were recruited from the general population, performed in the normal range on neuropsychological tests, had no clinical evidence of major neurological or psychiatric disorder, had a Mini-Mental State Examination (MMSE) ≥ 28, and were genotyped for apolipoprotein E (APOE) ε4 status. Amyloid PET scanning was added to the ongoing study in June 2010. The RNT assessment was completed between 1 September 2015 and 16 December 2017. Participants must have completed the RNT questionnaire and undergone at least one neuroimaging scan to be included in the analyses.

The IMAP/IMAP+ study was approved by the local ethics committee. Written informed consent was obtained from all participants.

### Psychological Measures

#### Repetitive Negative Thinking

The 15-item self-report Perseverative Thinking Questionnaire (PTQ)^23^ was translated into French by native speakers. Items use Likert scales ranging from 0 (never) to 4 (almost always), and all are positively scored with higher scores reflecting higher levels of RNT. Total scores range from 0-60. The PTQ was designed to measure content-independent levels of RNT, and has been validated for use in clinical and non-clinical populations.^23^

#### Depression

The Montgomery and Asberg Depression Rating Scale (MADRS) is a 10-item semi-structured interview that measures depressive symptoms. It has a maximum total score of 60, with higher scores indicating greater depressive symptomology.^24^

#### Anxiety

The trait subscale of the State-Trait Anxiety Inventory (STAI-B) is a 20-item self-report questionnaire that assesses habitual anxiety tendencies. Scores range from 20-80, with higher scores indicating greater anxiety symptomology.^25^

### Neuroimaging Acquisition and Processing

In this study, we used neuroimaging data acquired at baseline (T1), 18 months (T2) and 36 months (T3) after baseline, where available. All participants were scanned at Cyceron Center (Caen, France) on the same positron emission tomography (PET, Discovery RX VCT 64 PET-CT, General Electric Healthcare) and MRI (Philips Achievia 3.0T) scanners. Data were processed using the Statistical Parametric Mapping software (SPM, version 12, Wellcome Trust Centre for Neuroimaging, http://www.fil.ion.ucl.ac.uk/spm/software/spm12) as detailed elsewhere.^26,27^

Briefly, 10-minute late Amyvid® PET scans were acquired 50 minutes after intravenous injection of ≈4MBq/Kg of Florbetapir. For 18F-FDG-PET, after fasting for at least 6 hours and a 30-minute resting period, 180MBq of 18F-FDG was intravenously injected as a bolus. A 10-min PET acquisition scan began 50 minutes after injection.

PET data were corrected for partial volume effects (i.e. partial volume-corrected [PVC]) using the Muller-Gartner method,^28^ coregistered onto their corresponding MRI, and normalized using the deformation parameters defined from the MRI procedure. Resultant images were quantitatively normalized using a white matter reference region corresponding to the average of segmented white matter images from a group of healthy adults aged between 20 and 60 years, scanned at the same center, thresholded to 0.8 and eroded with a 6 mm^3^ sphere to avoid contamination by grey matter and cerebrospinal fluid binding (see ^27^ for details). SUVR values were extracted from resulting SUVR PET images for use in statistical analyses within predefined masks (see below).

T1-weighted images were segmented using multimodal segmentation (with Fluid Attenuated Inversion Recovery images), spatially normalized to the Montreal Neurological Institute (MNI) template, and modulated for nonlinear warping using the SPM12 Segment function so that resulting grey matter volume maps were corrected for brain size. Grey matter volume values were extracted from these images for use in statistical analyses within predefined masks (see below).

### Statistical Analyses

Associations between RNT, symptoms of depression and anxiety, and potential confounders (i.e. age at RNT assessment, sex, education, and APOE status) were investigated using separate correlation analyses for continuous variables and Student’s t-tests for categorical variables, with statistical significance set to P<0.05. APOE status was dichotomised into ɛ4-positive and ɛ4-negative.

The relationship between RNT, anxiety and depression and change in global cortical amyloid-PET (Florbetapir-PET), glucose metabolism (FDG-PET), structural grey matter volume over time was examined using separate linear mixed effects models. These models are robust to unbalanced and incomplete data in longitudinal designs^29^, and allowed for inclusion of all eligible data points for a given analysis. For PET images, PVC data were used in primary analyses.

#### Primary analyses

Primary analyses used amyloid-PET SUVR values extracted within the predefined global cortical mask^20^ from unsmoothed amyloid-PET images. FDG-PET SUVR values were extracted using the same procedure, within a predefined AD mask,^30^ from unsmoothed FDG-PET images.

A study-specific grey matter mask was generated from segmented images of all participants using SPM12. Mean tissue probability maps were calculated across the whole sample for grey matter, white matter, and cerebrospinal fluid. To define voxels predominantly corresponding to grey matter, voxels were retained when the grey matter probability was higher than both the white matter and cerebrospinal fluid probabilities, and when it exceeded 30%. The resulting mask was then applied to individual modulated, segmented and spatially normalized grey matter images in MNI space. Grey matter values were extracted voxel-wise for each participant from the grey matter volume maps corrected for brain size.

Regarding RNT, separate models with participant-level random intercepts, estimated via restricted maximum likelihood, were fitted for each neuroimaging outcome, with fixed effects of RNT, time (T1, T2, T3) and the interaction between RNT and time as the main explanatory variables. A random effect of individual was specified to account for repeated measures on participants over time. The model allowed both the baseline level of neuroimaging outcome and its rate of change over time to vary according to RNT level. Participant-specific baseline differences were accommodated by including a random intercept for participant. Model 1 included fixed effects of age at RNT assessment, sex, educational level, APOE status (when amyloid was the outcome variable). Model 2 included variables in Model 1, further adjusting for psychological symptoms (i.e., depressive and anxiety). Continuous variables were mean-centred. Similar models were fitted with either depression or anxiety and their respective interactions with time as the main explanatory variable, and the neuroimaging biomarker as the outcome. Model 2 was further adjusted for RNT and depressive/anxiety symptoms. The linearity of continuous predictors was evaluated by comparing the Model 1 with models including quadratic terms. No evidence of substantial departures from linearity was observed.

When significant associations were observed in the primary Model 2 analyses, post-hoc analyses were conducted by dividing the predictor variable (e.g. RNT) into tertiles to aid visualisation, and using Tukey’s honestly significant difference (HSD) test to control the family-wise error rate. Analyses were performed in R studio 2025.09.1+401.

Further, voxel-wise analyses were also conducted to identify regional changes in the neuroimaging outcome(s). PET images were smoothed using a 10-mm FWHM Gaussian kernel and grey matter volume maps with 8-mm FWHM Gaussian kernel. Linear mixed-effects models fitted at each voxel in R (version 4.4.2) were then performed with the lme4 package.^31^ All voxel-wise analyses were adjusted for age, sex, education and APOE status (when amyloid was the outcome variable), and psychological covariates. Results were evaluated for significance at false discovery rate (FDR)-corrected P<0.05 combined with a minimum cluster size determined by Monte-Carlo simulations using the AFNI’s 3dClustSim program to achieve a corrected statistical significance of P<0.05.

#### Sensitivity analyses

Sensitivity analyses were conducted in fully-adjusted models (Model 2) in restricted samples of participants aged **≥**50 and ≥60, without partial volume correction for amyloid-PET and FDG-PET outcomes and using neocortical grey matter masks (instead of predefined AD masks) for FDG-PET and grey matter volume outcomes.

## RESULTS

Participant characteristics are detailed in Table 1. RNT, depressive and anxiety symptoms were not associated with sex, education or APOE status (all *P*>0.05). Neither RNT nor anxiety symptoms were associated with age; however, depressive symptoms were negatively associated with age (r = -0.41, 95% CI -0.63 to -0.14, *P*<0.005). RNT was positively associated with depressive (r = 0.24, 95% CI 0.46 to 0.43, *P*=0.02) and anxiety symptoms (r = 0.5, 95% CI 0.33 to 0.63, *P*<0.001), and depressive and anxiety symptoms were positively correlated (r = 0.26, 95% CI 0.06 to 0.44, *P*=0.01).

**Table 1.**
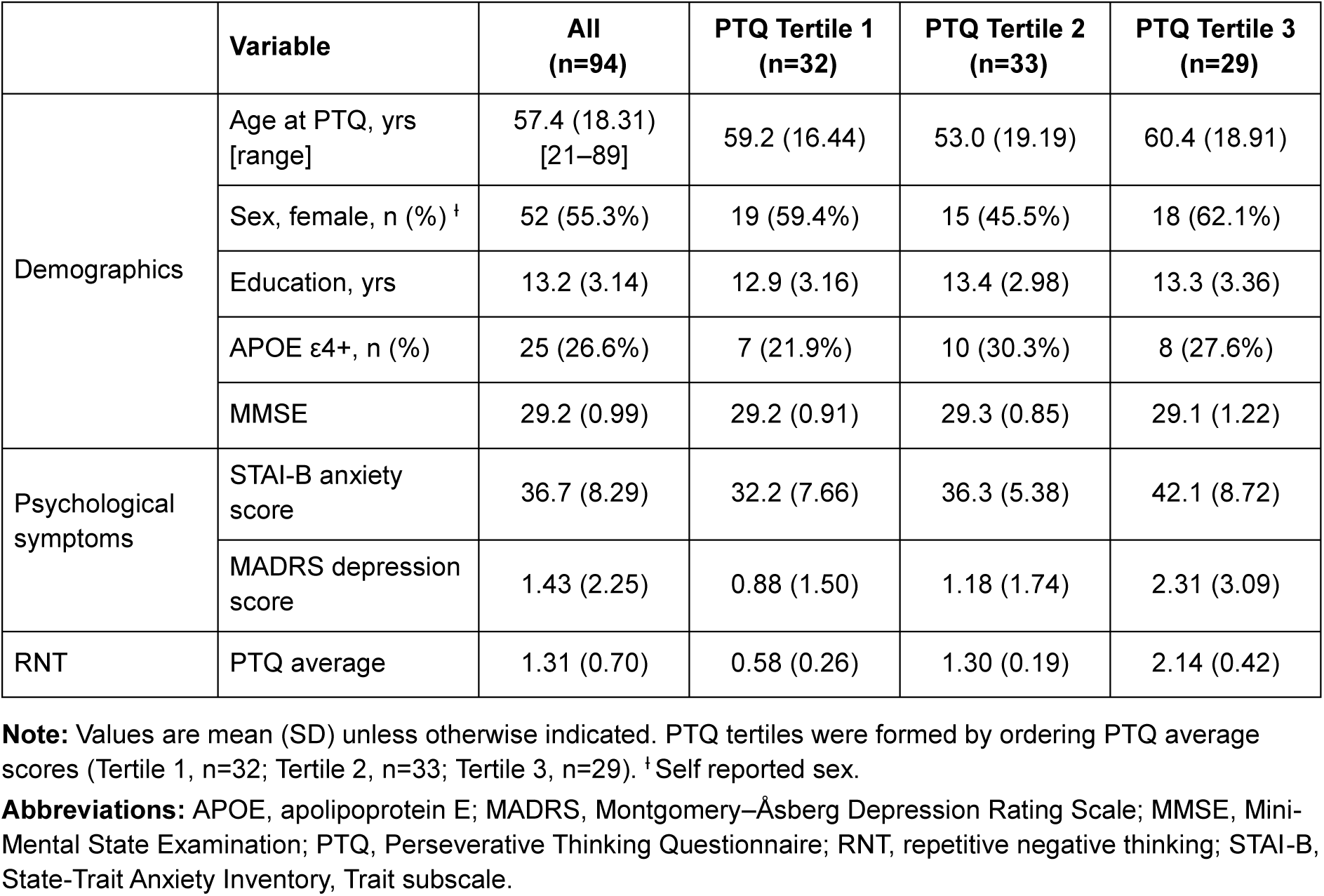
Participant Characteristics.

|  | **Variable** | **All<br>(n=94)** | **PTQ Tertile 1<br>(n=32)** | **PTQ Tertile 2<br>(n=33)** | **PTQ Tertile 3<br>(n=29)** |
|---|---|---|---|---|---|
| Demographics | Age at PTQ, yrs [range] | 57.4 (18.31)<br>[21–89] | 59.2 (16.44) | 53.0 (19.19) | 60.4 (18.91) |
|  | Sex, female, n (%) ǂ | 52 (55.3%) | 19 (59.4%) | 15 (45.5%) | 18 (62.1%) |
|  | Education, yrs | 13.2 (3.14) | 12.9 (3.16) | 13.4 (2.98) | 13.3 (3.36) |
|  | APOE ε4+, n (%) | 25 (26.6%) | 7 (21.9%) | 10 (30.3%) | 8 (27.6%) |
|  | MMSE | 29.2 (0.99) | 29.2 (0.91) | 29.3 (0.85) | 29.1 (1.22) |
| Psychological symptoms | STAI-B anxiety score | 36.7 (8.29) | 32.2 (7.66) | 36.3 (5.38) | 42.1 (8.72) |
|  | MADRS depression score | 1.43 (2.25) | 0.88 (1.50) | 1.18 (1.74) | 2.31 (3.09) |
| RNT | PTQ average | 1.31 (0.70) | 0.58 (0.26) | 1.30 (0.19) | 2.14 (0.42) |

**Note:** Values are mean (SD) unless otherwise indicated. PTQ tertiles were formed by ordering PTQ average scores (Tertile 1, n=32; Tertile 2, n=33; Tertile 3, n=29). ǂ Self reported sex.
**Abbreviations:** APOE, apolipoprotein E; MADRS, Montgomery–Åsberg Depression Rating Scale; MMSE, Mini-Mental State Examination; PTQ, Perseverative Thinking Questionnaire; RNT, repetitive negative thinking; STAI-B, State-Trait Anxiety Inventory, Trait subscale.

Of the total sample, global cortical amyloid-PET data were available for 46 (49%) at T1, 65 (69%) at T2, 23 (24%) at T3, resulting in 134 data points. FDG-PET data were available for 71 (76%) at T1, 90 (96%) at T2, 24 (26%) at T3, resulting in 185 data points. MRI data providing structural grey matter volume were available for 94 (100%) at T1, 91 (97%) at T2, 25 (27%) at T3, resulting in 210 data points. See Supplementary eTable 1 for details.

### Amyloid accumulation

We found evidence of faster amyloid accumulation over time with higher RNT in both Model 1 adjusting for confounders (Model 1: *B* = 0.0121; 95% CI 0.003 to 0.021, *P*=0.011;

Supplementary eTable 2) and Model 2 further adjusting for depressive and anxiety symptoms (Model 2: *B* = 0.0120, 95% CI 0.003 to 0.021, *P*=0.012; Table 2). Specifically, each one-point increase on the PTQ scale was associated with an additional 0.012-unit increase in amyloid SUVR per 18 months. Stratification by RNT tertiles showed that the estimated change in amyloid levels over 18 months was −0.0061 (95% CI −0.0185 to 0.0063) in the low, 0.0013 (95% CI −0.0123 to 0.0149) in the middle, and 0.0184 (95% CI 0.0089 to 0.0279) in the high RNT tertile groups. The rate of amyloid accumulation in the high RNT tertile was significantly greater than in the low tertile (difference in slopes = 0.0245, 95% CI 0.0057 to 0.0433, *P*=0.008; Figure 1), but did not significantly differ from the middle tertile (difference in slopes = 0.0171, 95% CI −0.0028 to 0.0370, *P*=0.105). There was no evidence of a difference in slopes between the low and middle tertiles (difference in slopes = −0.0074, 95% CI −0.0295 to 0.0147, *P*=0.703).

**Figure 1.**
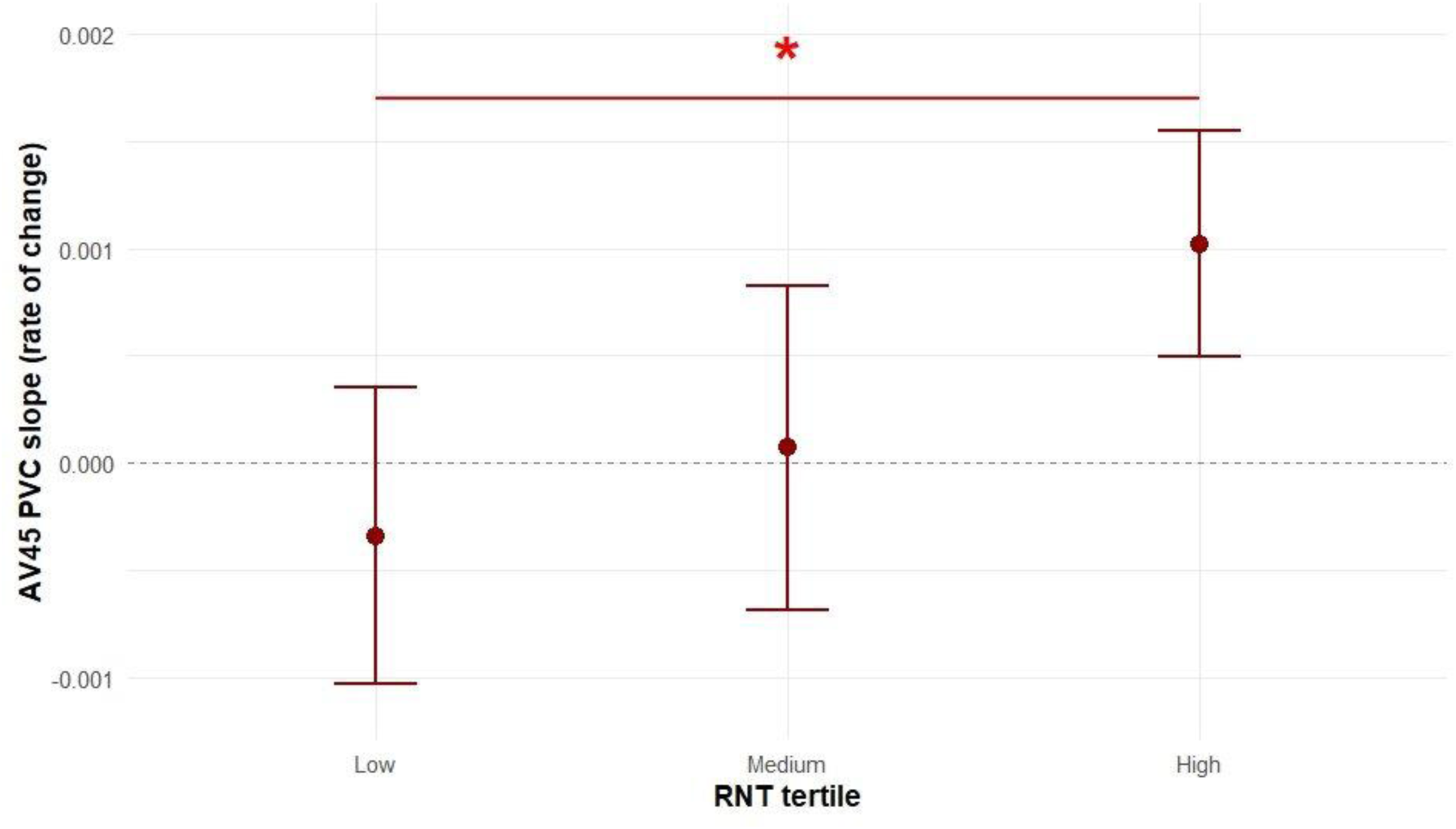
Association between tertile-stratified Repetitive Negative Thinking with longitudinal amyloid accumulation. Estimated rates of AV45 SUVR change by RNT tertile. Error bars indicate 95% CIs. The asterisk indicates a significant difference between the low and high RNT tertiles (Tukey-adjusted *P* = 0.003). Models were adjusted for age, sex, education, apolipoprotein E ε4 status, depressive and anxiety symptoms. Abbreviations: CI, confidence interval; RNT, repetitive negative thinking.

**Table 2.** Cross-sectional and longitudinal relationships between (a) RNT and AD biomarkers, (b) depressive symptoms and AD biomarkers, (c) anxiety symptoms and AD biomarkers in fully adjusted models.

| **Variable** | **Amyloid<sup>a</sup>** |  | **Glucose metabolism<sup>b</sup>** |  | **Grey matter volume<sup>c</sup>** |  |
|---|---|---|---|---|---|---|
|  | **Est. (95% CI)** | ***P*** | **Est. (95% CI)** | ***P*** | **Est. (95% CI)** | ***P*** |
| **(a) RNT<sup>1</sup>** |  |  |  |  |  |  |
| RNT | -0.0157 (-0.048, 0.016) | 0.339 | -0.0022 (-0.061, 0.056) | 0.941 | -0.0029 (-0.012, 0.006) | 0.533 |
| Time | 0.0059 (-0.001, 0.013) | 0.089 | -0.0057 (-0.024, 0.013) | 0.549 | -0.0025 (-0.004, -0.001) | 0.005 |
| RNT × Time | 0.0120 (0.003, 0.021) | 0.011 | 0.0004 (-0.027, 0.028) | 0.979 | 0.0014 (-0.001, 0.004) | 0.283 |
| **(b) Depressive symptoms<sup>2</sup>** |  |  |  |  |  |  |
| Depressive symptoms | -0.0011 (-0.009, 0.007) | 0.803 | -0.0005 (-0.017, 0.016) | 0.950 | -0.0009 (-0.004, 0.002) | 0.508 |
| Time | 0.0072 (0.000, 0.014) | 0.041 | -0.0057 (-0.024, 0.013) | 0.549 | -0.0024 (-0.004, -0.001) | 0.008 |
| Depressive symptoms × Time | 0.0022 (-0.001, 0.006) | 0.184 | -0.0016 (-0.011, 0.007) | 0.727 | 0.0008 (-0.000, 0.002) | 0.088 |
| **(c) Anxiety symptoms<sup>3</sup>** |  |  |  |  |  |  |
| Anxiety symptoms | -0.0008 (-0.003, 0.002) | 0.566 | 0.0004 (-0.004, 0.005) | 0.875 | -0.0003 (-0.001, 0.000) | 0.487 |
| Time | 0.0062 (-0.001, 0.013) | 0.082 | -0.0056 (-0.025, 0.013) | 0.563 | -0.0026 (-0.004, -0.001) | 0.005 |
| Anxiety symptoms × Time | 0.0006 (-0.000, 0.001) | 0.091 | 0.0000 (-0.002, 0.002) | 0.978 | 0.0001 (-0.000, 0.000) | 0.536 |

All estimates are adjusted for age, sex, education, and apolipoprotein E ε4 status (for amyloid analyses). <sup>1</sup> Additionally adjusted for depressive and anxiety symptoms; <sup>2</sup> additionally adjusted for RNT and anxiety symptoms; <sup>3</sup> additionally adjusted for RNT and depressive symptoms.
**Note:** All continuous estimates, apart from time, are mean-centred. Time indicates an 18-month interval.
<sup>a</sup> Partial volume corrected. Participants, N = 76; observations, N = 134. <sup>b</sup> Partial volume corrected using an AD mask. Participants, N = 94; observations, N = 185. <sup>c</sup> AD mask. Participants, N = 95; observations, N = 210.
Abbreviations: AD, Alzheimer disease; APOE, apolipoprotein E; CI, confidence interval; RNT, repetitive negative thinking.

The post-hoc FDR-corrected voxel-wise linear mixed-effects model showed that RNT predicted faster Aβ accumulation within posterior cingulate and precuneus areas, as well as the frontal cortex including medial frontal areas and more lateral frontal regions, together with the temporo-parietal cortex and superior parietal region (*B* = 0.021, 95% CI 0.012 to 0.03, *P*<0.001; Figure 2).

**Figure 2.**
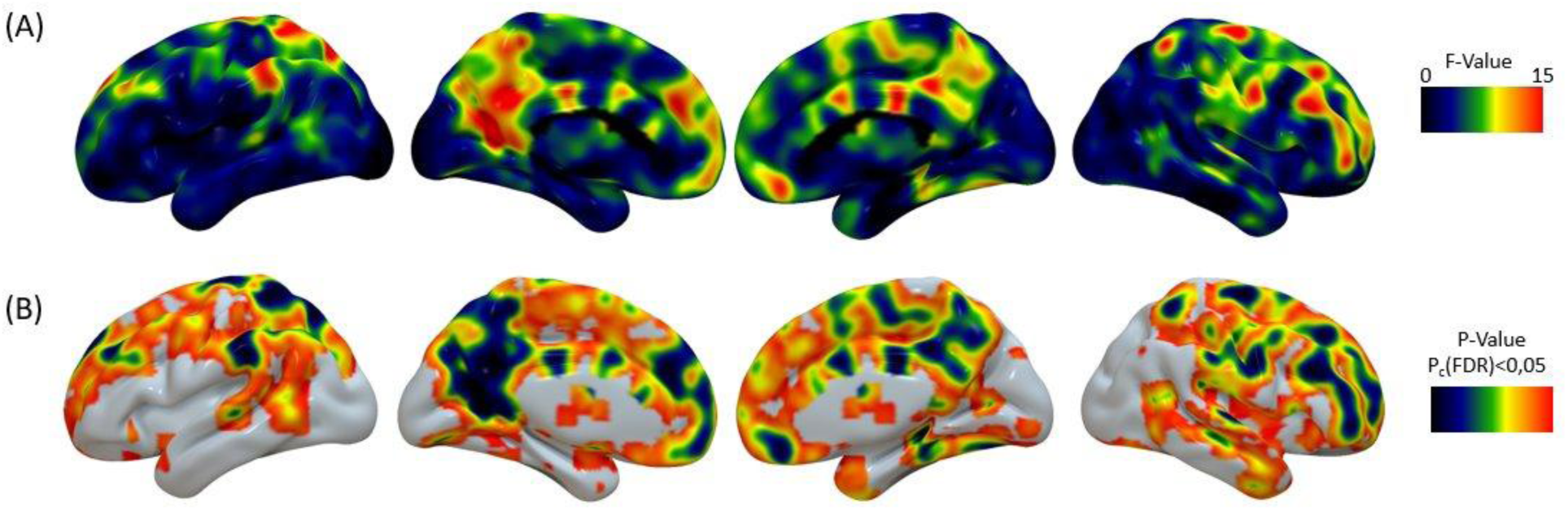
Voxel-wise association between repetitive negative thinking and longitudinal change in amyloid deposition. Results of voxel-wise linear mixed-effects models testing the interaction between RNT (PTQ score) and time on grey matter Flobetapir SUVR. Models included a participant-level random intercept and were adjusted for age, sex, education, APOE ε4 status, and depressive and anxiety symptom (Model 2). (A) Unthresholded map of F-statistic for the RNT + time interaction. (B) Map restricted to voxels surviving FDR correction (P < 0.05).

Results remained consistent in sensitivity analyses restricting participants to those aged **≥**50 and ≥60; and were slightly attenuated when using masks without partial-volume correction (Supplementary eTable 3).

We found no evidence that depressive or anxiety symptoms were associated with rate of change in amyloid accumulation in either model or in any sensitivity analyses (Table 2, Supplementary eTable 2 and 3).

### Glucose metabolism

We found no evidence that RNT, depressive or anxiety symptoms were associated with longitudinal change in glucose metabolism in either model or in any sensitivity analyses (*P*s>0.05, Table 2, Supplementary eTable 2 and 4).

### Grey matter atrophy

We found no evidence that RNT, depressive or anxiety symptoms were associated with longitudinal change in grey matter volume in either model or in any sensitivity analyses (*P*s>0.05, Table 2, Supplementary eTable 2 and 5).

## DISCUSSION

The current study sought to determine whether RNT was associated with change in established neurobiological markers of AD over time in cognitively healthy adults. We found that higher levels of RNT were associated with more rapid accumulation of amyloid, but not with change in glucose metabolism or grey matter volume, over a 3-year period. Neither depressive nor anxiety symptoms were associated with change in any biomarker.

RNT is a cognitive-emotional process that influences the development and perpetuation of psychological risk factors for dementia, including depression and anxiety.^32^ In our study, RNT was associated with greater amyloid accumulation in brain regions vulnerable to early AD-related amyloid aggregation, even after accounting for known predictors of amyloid deposition — age, APOE ɛ4 status — and of AD risk – sex, education, depressive and anxiety symptoms. Indeed while RNT was also associated with symptoms of depression and anxiety, neither of these symptoms were themselves associated with amyloid accumulation, indicating specificity of this thinking style with neuropathological change. These findings add to accumulating evidence implicating RNT as a marker of AD pathogenesis.^10,11,13,14^

We had previously proposed that RNT may increase AD risk through the stress pathway, given its association with acute changes in indicators of stress (e.g., cortisol, elevated blood pressure, etc)^33^ and associations between stress, cognitive decline, clinical AD and AD biomarkers.^34,35^ RNT may perpetuate the cognitive and neural response to stressful experiences. Neuroimaging evidence from both resting state and task-based functional connectivity studies indicate RNT activation of brain regions in the default mode, salience and executive control networks – implicated in self-referential and emotional processing, and cognitive control.^36^ Subsequent network-level work has associated RNT with dysregulated system segregation of these same networks, while experimental evidence indicates that negative thinking is associated with lasting alterations in posterior default mode network–limbic connectivity following negative socio-emotional experiences. Further, high neuronal activity has been linked with early amyloid accumulation.^37^ Based on these findings, we cautiously suggest that RNT may lead to a failure to disengage and recover from stressful experiences resulting in persistent and dysregulated neural activation and connectivity, which may increase vulnerability to early amyloid accumulation. However the reverse may also be the case – early amyloid accumulation could disrupt neural circuitry leading to higher RNT.

Our failure to observe associations between RNT and change in glucose metabolism or grey matter atrophy may be due to the relatively young age and health of the sample, as amyloid accumulation is believed to be the earliest AD-related pathological change.^38^ Indeed we and others have previously observed RNT associations with markers of atrophy in older samples;^17,18^ however in this study our null cross-sectional and longitudinal results remained even after restricting the sample to older adults. These previous studies examined plasma-derived neurofilament light, a cytoskeletal protein released after axonal damage or neuronal degeneration,^39^ and a machine learning-derived estimate of brain age, suggesting that associations may depend on the type and sensitivity of the pathological marker.

### Limitations

Neuroimaging data were often unavailable, especially for amyloid. It is important to note that these data were largely unavailable due to visits not being conducted, rather than participant dropout, therefore were less likely to be influenced by survivor bias. However, we cannot be sure that the missing data did not bias estimates of magnitude and direction of amyloid trajectories. We sought to mitigate this possibility by using multi-level models of longitudinal data and including factors associated with missingness as covariates. RNT assessments coincided with different neuroimaging time points allowing the possibility of reverse causation – emerging amyloid pathology could have influenced RNT levels. It also relies on the assumption that RNT levels are largely constant, an assumption supported by our previous finding that RNT is relatively stable over at least a 1-year interval using the same measure.^11^ Finally, the means and variances in depression and anxiety scores were relatively low. While a population with greater depression and anxiety symptomology may have exposed associations with amyloid accumulation, recent systematic reviews and meta-analyses reported no cross-sectional associations between amyloid and anxiety^40^ or depression^41^ in cognitively healthy adults; thus offering limited support for that possibility at present.

### Future

Work leveraging nationally provided psychological therapy records linked with medical records in England suggest that reliable improvements in depression and/or anxiety with psychological therapy are associated with reduced risk of dementia up to eight years later.^42,43^ Additionally, RNT is a key mediator of change in depression and anxiety following psychological therapy,^44^ suggesting that the reduced dementia risk observed may act via reductions in RNT. Future research could directly investigate whether a) RNT can be reduced in older and/or at-risk adults, and b) reductions in RNT through intervention are associated with change in AD markers. Further, despite the dynamic nature of thoughts, clinical research still relies on static, retrospective, and self-reported assessments.^45^ integrating advances in ecological momentary assessment, artificial intelligence and large language models to categorise thoughts could help determine which aspect(s) of RNT are most pernicious and inform targeted risk-reduction interventions.

## Conclusion

We observed an association between RNT and amyloid accumulation over three years in cognitive healthy adults. Collectively, the accumulating behavioural and neurobiological evidence implicates RNT as an early marker and/or driver of AD pathology.

## Data Availability

Data related to the current study are derived from the IMAP/IMAP+ study. The datasets used and/or analyzed during the current study are available to research groups on reasonable request and pending approval by the study coordinator.

## ACKNOWLEDGEMENTS

Marchant received funding from the University of Caen Normandy (UCN) Visiting Professor program. Independent of this work, Marchant has received consulting fees from Roche. The IMAP/IMAP+ study was supported by Fondation Plan Alzheimer (Alzheimer Plan 2008-2012); Programme Hospitalier de Recherche Clinique (PHRCN 2011-A01493-38 and PHRCN 2012 12-006-0347); Agence Nationale de la Recherche (LONGVIE 2007); Région Basse-Normandie; Association France Alzheimer et maladies apparentées AAP 2013. The authors would like to thank the people who contributed to data acquisition and/or processing, the Cyceron staff members and the volunteers who were included in this study.

Dr Marchant had full access to all the data in the study and takes responsibility for the integrity of the data and the accuracy of the data analysis. Funding sources had no role in the design and conduct of the study; collection, management, analysis, and interpretation of the data; preparation, review, or approval of the manuscript; and the decision to submit the manuscript for publication.

## DATA SHARING STATEMENT

Data related to the current study are derived from the IMAP/IMAP+ study. The datasets used and/or analysed during the current study are available to research groups on reasonable request and pending approval by the study coordinator.

## AUTHOR CONTRIBUTIONS

*Study concept and design:* Marchant, Chételat.

*Acquisition, analysis, or interpretation of data:* All authors.

*Drafting of the manuscript:* Marchant.

*Critical revision of the manuscript for important intellectual content:* All authors.

*Obtained funding:* Marchant, Chételat.

*Administrative, technical, or material support:* Mezenge.

## References

1. Frisoni GB, Hansson O, Nichols E, et al. New landscape of the diagnosis of Alzheimer’s disease. The Lancet. 2025;406(10510):1389–1407. doi:10.1016/S0140-6736(25)01294-2

2. Beason-Held LL, Goh JO, An Y, et al. Changes in Brain Function Occur Years before the Onset of Cognitive Impairment. J Neurosci. 2013;33(46):18008–18014. doi:10.1523/JNEUROSCI.1402-13.2013

3. Brain J, Alshahrani M, Kafadar AH, et al. Temporal dynamics in the association between depression and dementia: an umbrella review and meta-analysis. eClinicalMedicine. 2025;84. doi:10.1016/j.eclinm.2025.103266

4. Santabarbara J, Lipnicki DM, Bueno-Notivol J, Olaya-Guzman B, Villagrasa B, Lopez-Anton R. Updating the evidence for an association between anxiety and risk of Alzheimer’s disease: A meta-analysis of prospective cohort studies. J Affect Disord. 2020;262:397–404. doi:10.1016/j.jad.2019.11.065

5. Marchant NL, Howard RJ. Cognitive Debt and Alzheimer’s Disease. Journal of Alzheimer’s Disease. 2015;44(3):755–770. doi:10.3233/JAD-141515

6. Marchant NL, Bartres Faz D, Stott J, Terracciano A, Chételat G. Psychological risk, protection and support in the context of dementia. Nat Mental Health. Published online October 23, 2026. doi:10.1038/s44220-026-00748-9

7. Harvey AG, Watkins E, Mansell W. Cognitive Behavioural Processes across Psychological Disorders: A Transdiagnostic Approach to Research and Treatment. Oxford University Press, USA; 2004.

8. Morse RM, Koutsoubelis F, Whitfield T, et al. Worry and ruminative brooding: associations with cognitive and physical health in older adults. Front Psychol. 2024;15. doi:10.3389/fpsyg.2024.1332398

9. Schlosser M, Demnitz-King H, Whitfield T, Wirth M, Marchant NL. Repetitive negative thinking is associated with subjective cognitive decline in older adults: a cross-sectional study. BMC Psychiatry. 2020;20(1):500. doi:10.1186/s12888-020-02884-7

10. Ye N, Peng L, Deng B, et al. Repetitive negative thinking is associated with cognitive function decline in older adults: a cross-sectional study. BMC Psychiatry. 2025;25(1):562. doi:10.1186/s12888-025-06815-2

11. Marchant NL, Lovland LR, Jones R, et al. Repetitive negative thinking is associated with amyloid, tau, and cognitive decline. Alzheimer’s & Dementia. 2020;16(7):1054–1064. doi:10.1002/alz.12116

12. Pietrzak RH, Maruff P, Woodward M, et al. Mild worry symptoms predict decline in learning and memory in healthy older adults: A 2-year prospective cohort study. Am J Geriatr Psychiatry. 2012;20(3):266–275.

13. Stephan Y, Sutin AR, Mõttus R, Luchetti M, Aschwanden D, Terracciano A. Personality nuances and risk of dementia: Evidence from two longitudinal studies. Journal of Psychiatric Research. 2024;175:1–8. doi:10.1016/j.jpsychires.2024.04.039

14. Lee S, Zide BS, Palm ST, et al. Specific Association of Worry With Amyloid-β But Not Tau in Cognitively Unimpaired Older Adults. The American Journal of Geriatric Psychiatry. 2024;32(10):1203–1214. doi:10.1016/j.jagp.2024.04.016

15. Schwarz C, Lange C, Benson GS, et al. Severity of Subjective Cognitive Complaints and Worries in Older Adults Are Associated With Cerebral Amyloid-β Load. Front Aging Neurosci. 2021;13:675583. doi:10.3389/fnagi.2021.675583

16. Boscheck H, Luft M, Mauer R, et al. Pathways to resilience: relationships between cognitive reserve, psychological debt, and Alzheimer’s disease biomarkers. Alz Res Therapy. 2026;18(1):93. doi:10.1186/s13195-026-02054-z

17. Lau Y, Bansal A, Palix C, et al. Sex differences in the association between repetitive negative thinking and neurofilament light. npj Mental Health Res. 2024;3(1):53. doi:10.1038/s44184-024-00093-8

18. Karim HT, Ly M, Yu G, et al. Aging faster: worry and rumination in late life are associated with greater brain age. Neurobiol Aging. 2021;101:13–21. doi:10.1016/j.neurobiolaging.2021.01.009

19. Schwarz C, Benson GS, Daria A, et al. Negative affective burden is associated with higher resting-state functional connectivity in subjective cognitive decline. Scientific Reports (Nature Publisher Group). 2022;12(1). doi:10.1038/s41598-022-10179-y

20. La Joie R, Perrotin A, Barré L, et al. Region-Specific Hierarchy between Atrophy, Hypometabolism, and β-Amyloid (Aβ) Load in Alzheimer’s Disease Dementia. J Neurosci. 2012;32(46):16265–16273. doi:10.1523/JNEUROSCI.2170-12.2012

21. Perrotin A, de Flores R, Lamberton F, et al. Hippocampal Subfield Volumetry and 3D Surface Mapping in Subjective Cognitive Decline. J Alzheimers Dis. 2015;48 Suppl 1:S141–50. doi:10.3233/jad-150087

22. Perrotin A, La Joie R, de La Sayette V, et al. Subjective cognitive decline in cognitively normal elders from the community or from a memory clinic: Differential affective and imaging correlates. Alzheimers Dement. 2017;13(5):550–560. doi:10.1016/j.jalz.2016.08.011

23. Ehring T, Zetsche U, Weidacker K, Wahl K, Schonfeld S, Ehlers A. The Perseverative Thinking Questionnaire (PTQ): validation of a content-independent measure of repetitive negative thinking. J Behav Ther Exp Psychiatry. 2011;42(2):225–232. doi:10.1016/j.jbtep.2010.12.003

24. Montgomery SA, Åsberg M. A New Depression Scale Designed to be Sensitive to Change. Br J Psychiatry. 1979;134(4):382–389. doi:10.1192/bjp.134.4.382

25. Spielberger CD, Gorsuch RL, Lushene R, Vagg PR, Jacobs GA. Manual for the State-Trait Anxiety Inventory. Consulting Psychologists Press; 1983.

26. Kuhn E, Moulinet I, Perrotin A, et al. Cross-sectional and longitudinal characterization of SCD patients recruited from the community versus from a memory clinic: subjective cognitive decline, psychoaffective factors, cognitive performances, and atrophy progression over time. Alz Res Therapy. 2019;11(1):61. doi:10.1186/s13195-019-0514-z

27. Gonneaud J, Arenaza-Urquijo EM, Mézenge F, et al. Increased florbetapir binding in the temporal neocortex from age 20 to 60 years. Neurology. 2017;89(24):2438–2446. doi:10.1212/WNL.0000000000004733

28. Müller-Gärtner HW, Links JM, Prince JL, et al. Measurement of radiotracer concentration in brain gray matter using positron emission tomography: MRI-based correction for partial volume effects. J Cereb Blood Flow Metab. 1992;12(4):571–583. doi:10.1038/jcbfm.1992.81

29. Pinheiro JC, Bates DM. Mixed-Effects Models in S and S-PLUS. Springer; 2000.

30. Besson FL, La Joie R, Doeuvre L, et al. Cognitive and Brain Profiles Associated with Current Neuroimaging Biomarkers of Preclinical Alzheimer’s Disease. J Neurosci. 2015;35(29):10402–10411. doi:10.1523/JNEUROSCI.0150-15.2015

31. Bates D, Mächler M, Bolker B, Walker S. Fitting Linear Mixed-Effects Models Using **lme4**. J Stat Soft. 2015;67(1). doi:10.18637/jss.v067.i01

32. Spinhoven P, Van Hemert AM, Penninx BW. Repetitive negative thinking as a predictor of depression and anxiety: A longitudinal cohort study. Journal of Affective Disorders. 2018;241:216–225. doi:10.1016/j.jad.2018.08.037

33. Ottaviani C, Thayer JF, Verkuil B, et al. Physiological concomitants of perseverative cognition: A systematic review and meta-analysis. Psychological Bulletin. 2016;142:231–259. doi:10.1037/bul0000036

34. Ouanes S, Popp J. High Cortisol and the Risk of Dementia and Alzheimer’s Disease: A Review of the Literature. Frontiers in Aging Neuroscience. 2019;Volume 11-2019. https://www.frontiersin.org/journals/aging-neuroscience/articles/10.3389/fnagi.2019.00043

35. Seeman TE, Singer BH, Rowe JW, Horwitz RI, McEwen BS. Price of adaptation--allostatic load and its health consequences. MacArthur studies of successful aging. Arch Intern Med. 1997;157(19):2259–2268.

36. Makovac E, Fagioli S, Rae CL, Critchley HD, Ottaviani C. Can’t get it off my brain: Meta-analysis of neuroimaging studies on perseverative cognition. Psychiatry Res Neuroimaging. 2020;295:111020. doi:10.1016/j.pscychresns.2019.111020

37. Bero AW, Yan P, Roh JH, et al. Neuronal activity regulates the regional vulnerability to amyloid-β deposition. Nat Neurosci. 2011;14(6):750–756. doi:10.1038/nn.2801

38. Hampel H, Hardy J, Blennow K, et al. The Amyloid-β Pathway in Alzheimer’s Disease. Mol Psychiatry. 2021;26(10):5481–5503. doi:10.1038/s41380-021-01249-0

39. Gaiottino J, Norgren N, Dobson R, et al. Increased Neurofilament Light Chain Blood Levels in Neurodegenerative Neurological Diseases. Reindl M, ed. PLoS ONE. 2013;8(9):e75091. doi:10.1371/journal.pone.0075091

40. Demnitz-King H, Saba L, Lau Y, et al. Association between anxiety symptoms and Alzheimer’s disease biomarkers in cognitively healthy adults: A systematic review and meta-analysis. Journal of Psychosomatic Research. 2023;166:111159. doi:10.1016/j.jpsychores.2023.111159

41. Twait EL, Wu JH, Kamarioti M, et al. Association of amyloid-beta with depression or depressive symptoms in older adults without dementia: a systematic review and meta-analysis. Transl Psychiatry. 2024;14(1):25. doi:10.1038/s41398-024-02739-9

42. John A, Saunders R, Desai R, et al. Associations between psychological therapy outcomes for depression and incidence of dementia. Psychol Med. 2023;53(11):4869–4879. doi:10.1017/S0033291722002537

43. Stott J, Saunders R, Desai R, et al. Associations between psychological intervention for anxiety disorders and risk of dementia: a prospective cohort study using national health-care records data in England. The Lancet Healthy Longevity. 2023;4(1):e12–e22. doi:10.1016/S2666-7568(22)00242-2

44. Gu J, Strauss C, Bond R, Cavanagh K. How do mindfulness-based cognitive therapy and mindfulness-based stress reduction improve mental health and wellbeing? A systematic review and meta-analysis of mediation studies. Clin Psychol Rev. 2015;37:1–12. doi:10.1016/j.cpr.2015.01.006

45. Verkuil B, Brosschot JF, Thayer JF. Capturing worry in daily life: Are trait questionnaires sufficient? Behaviour Research and Therapy. 2007;45(8):1835–1844. doi:10.1016/j.brat.2007.02.004

